# Cohort profile: The Nepal Turnaway Study

**DOI:** 10.64898/2026.03.10.26348060

**Authors:** Rachel Murro, Sarah Raifman, W John Boscardin, Mahesh C Puri, Sunita Karki, Anupama Ale Magar, Dev Chandra Maharjan, Corinne H Rocca, M Antonia Biggs, Nadia G Diamond-Smith, Diana Greene Foster

## Abstract

**Purpose:** The Nepal Turnaway Study was designed to understand abortion care experiences and the longitudinal wellbeing of abortion-seekers and their families.

**Participants:** The Nepal Turnaway Study is a nation-wide cohort of abortion seekers recruited from public and private facilities across all seven provinces between April 2019 and December 2020. It contains 1,832 abortion seekers followed for up to five years. Repeated measures of socioeconomic status, health and wellbeing—including maternal physical and mental health and child health (under age three)—were collected every six months or annually, alongside detailed pregnancy and abortion-seeking histories.

**Findings to date:** Abortion seekers in this context can be recruited at health facilities (96% participation) and successfully followed for up to five years (85% retention). Nearly half (49%, n=856) of abortion seekers were initially denied their abortion, and 16% (n=275) ultimately carried the pregnancy to term and gave birth. Those who were denied were more likely to be socioeconomically disadvantaged prior to abortion seeking.

**Future plans:** The Nepal Turnaway Study will be used to understand the longitudinal health and socioeconomic effects of receiving a wanted abortion in this setting. Exposure-balancing weights can be applied to ensure rigorous estimation of the effects of abortion denial on longitudinal outcomes. This cohort can also be used more broadly to examine the trajectories of women and their families in the years following abortion-seeking.

**Registration:** This study has been registered at clinicaltrials.gov (NCT03930576)

## INTRODUCTION

Globally, over 121 million unintended pregnancies occur each year.[1] Unwanted or mistimed births can be associated with negative physical, social and economic consequences; [2,3] the ability to avoid and terminate unintended pregnancies is thus a fundamental component of reproductive health and autonomy. However, in low-income countries, 60% of unintended pregnancies do not end in abortion,[1] and over three quarters of abortions are performed in unsafe conditions.[4] Carrying an unwanted pregnancy to term may lead to poor health outcomes for both birthing people and their families,[5] and the consequences of unsafe abortion can include infection, infertility, and death.[6] An estimated eight percent of maternal deaths worldwide are due to unsafe abortion,[7] though this is likely an underestimate due to the difficulty of measuring abortion-related mortality and morbidity.[7,8]

The Government of Nepal has taken steps to protect options for fertility planning as part of women’s reproductive health services. In 2002, abortion was legalized under certain circumstances by amending the country’s Penal Code (Mulki Ain)[9] and in 2018, parliament passed a separate measure, The Safe Motherhood and Reproductive Health Rights (SMRHR) Act, which guaranteed women’s rights to legal and safe abortion care on wider grounds.[10] Under the SMRHR Act, abortion is permitted with the consent of the pregnant woman up to 12 weeks’ gestation and up to 28 weeks’ in cases of rape, incest or the following situations: if the pregnant woman is living with HIV or some other incurable disease; if the pregnancy poses a danger to the woman’s life, physical health or mental health; or if there is a fetal anomaly. These national efforts are reflective of a significant shift in women rights and health. Prior to 2002, many women were imprisoned for having abortions.[11] Between 2004 and 2014, national laws and policies have enabled an estimated 850,000 women in Nepal to obtain safe, legal services.[12]

Despite national efforts to expand access to legal and safe abortion services over the past 25 years, many Nepali women, especially those living in poverty and who are geographically isolated from healthcare systems, remain unable to get care.[13,14] Notably, women who live in rural areas (over 80% of the country’s population) report substantial challenges in accessing services.[14] Under current law, only physicians and midlevel providers certified in safe abortion care by the government are eligible to provide induced abortion services, limiting service delivery.[15] Further, stigma and insufficient knowledge of the law prevent women from obtaining care, even if they are eligible for abortion under the law and it is theoretically affordable.[16,17] Poor availability of and access to legal services may lead women to 1) seek unsafe alternatives or 2) carry an unwanted pregnancy to term. As of 2014, over half of abortions in Nepal were performed without a certified provider or in other illegal circumstances.[18] The burden of unwanted births due to lack of abortion access in Nepal remains understudied.

### Study aims

A team of researchers from the University of California, San Francisco in the United States and CREHPA in Kathmandu, Nepal collaborated to adapt the successful, prospective cohort design of the Turnaway Study in the U.S. to Nepal. The U.S. Turnaway Study evaluated the mental health, physical health, and socioeconomic outcomes of receiving an abortion compared to carrying an unwanted pregnancy to term.

The Nepal Turnaway Study is a prospective longitudinal study of women with unwanted pregnancies – pregnancies which they seek to abort rather than carry to term – designed chiefly to investigate the consequences for women and their families of legal abortion, abortion outside of the legal system, and delivery of an unwanted pregnancy in Nepal. The objective of this paper is to describe the Nepal Turnaway Study and invite collaboration from those interested in using the underlying data for novel research questions.

## METHODS

### Recruitment and enrollment

From April 2019 through December 2020, women seeking abortion care were recruited at 22 facilities across Nepal (Fig 1). Initial recruitment began at 14 public and private/non-profit sites, with one of each type selected in all seven provinces. Facilities were chosen randomly, with the probability of selection proportional to client volume, from the national list of certified abortion facilities that had performed at least 60 abortions per year during 2016–2017. These sites together accounted for most legal abortion services provided nationally at that time. In late 2019, seven of the original facilities were replaced and one new site was added because of limited numbers of eligible participants at seven locations. The same sampling approach was used to identify the new site. Recruitment was suspended twice during the study period: first between October 2019 and January 2020 to replace the seven facilities with low numbers of eligible women, and again from April to June 2020 because of travel restrictions associated with the COVID-19 pandemic.

**Figure 1.**
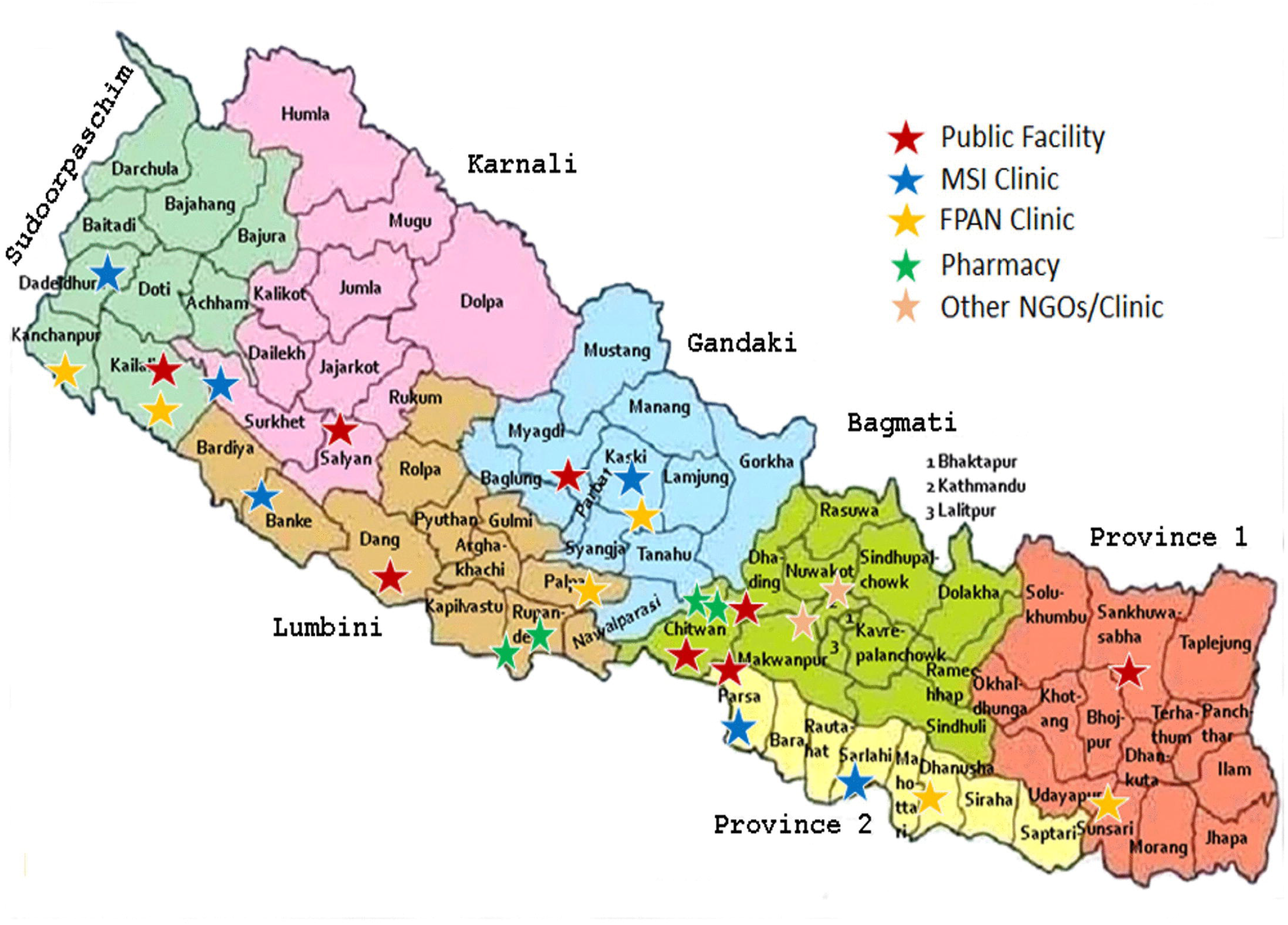
Map of Nepal with Turnaway study sites.

Women aged 15 years and older who sought abortion care at participating study facilities and resided in Nepal were eligible for enrollment. In the initial recruitment phase (April 15–May 10, 2019), women at any gestational age were invited to participate. From May 11, 2019 through December 31, 2020, eligibility was limited to women presenting at or beyond ten weeks of gestation, or those uncertain of their gestational age, to ensure adequate representation of individuals more likely to be denied services. No additional exclusion criteria were applied.

100% of abortion seekers were screened for study eligibility by a doctor, nurse, or counselor upon arrival to the facility. These point people recorded the patient’s age and estimated gestation (reported by the patient based on their last menstrual period), determined the patient’s eligibility for legal abortion, and recorded the reason for abortion ineligibility, if relevant. If a woman was eligible for the study, she was subsequently taken to a private clinic room to speak with a trained research staff member. The research staff member confirmed study eligibility and obtained written informed consent. A thumbprint was obtained for women unable to sign. In the case of minors under 18 years of age, participants provided assent for participation and interviewers obtained consent from one biological parent.

Ethical review and approval was granted by The University of California, San Francisco Human Research Protection Program (18-25863) and the Ethics Committee of the Nepal Health Research Council (704/2018).

### Data collection procedures

After obtaining informed consent and confirming eligibility, the research staff member then conducted the baseline survey, which included collection of sociodemographic characteristics. All participants completed the baseline survey prior to obtaining abortion services. Subsequent interviews took place six weeks after the initial visit and then at six-month intervals over the next five years at the participant’s home or another chosen place, covering all health and economic outcomes measurement. Interviews were conducted in Nepali, Maithali, Tharu, Bhojpuri, or Hindi, according to the participant’s preference and took about 40 minutes. Participants received financial compensation of NRs 500 (approximately $4 USD) for the baseline and each subsequent interview. Research staff members conducted each interview using a tablet and uploaded survey answers to a secure web-based storage platform. Participants initially consented to participate for up to three years. In an effort to maximize the period of observations, we put resources toward gathering longer term data, beginning in March 2023. At that point, consent was obtained for all willing participants to continue surveys each six months for up to five years. We ceased interviews when funding was exhausted in December 2024, at which point some participants had completed five years of interviews and others were administratively censored.

### Measures

The Nepal Turnaway study collected data on a range of measures to capture the social, economic, emotional, mental, and physical wellbeing of participants and their families. Table 1 documents all measures and collection frequency.

**Table 1.** List of survey measures.

| Category | Indicators | BL | 6w | 6m | 12m | 18m | 24m | 30m | 36m | 42m | 48m | 54m | 60m |
| --- | --- | --- | --- | --- | --- | --- | --- | --- | --- | --- | --- | --- | --- |
| Respondent demographics<sup>a</sup> | Schooling, student status, work status/type; marriage/cohabitation status; household composition; household income adequacy; caste/ethnicity; religion; parity (lifetime history of pregnancies, births, miscarriages, abortions) | ? | ? | ? | ? | ? | ? | ? | ? | ? | ? | ? | ? |
| Life goals | Goals (school, work, marriage, childbearing, other) for next one year | ? |  |  | ? |  | ? |  |  |  |  | ? |  |
| Life goals progress | Reflect whether 1-year goals from year prior were achieved |  |  |  | ? |  | ? |  | ? |  |  |  |  |
| Husband demographics<sup>b</sup> | Husband age; husband schooling; age at cohabitation; marriage type; dowry |  | ? | ? | ? | ? | ? | ? | ? | ? | ? | ? | ? |
| Husband employment & migration | Husband work status/type; husband work abroad; current living arrangement/location |  | ? | ? | ? | ? | ? | ? | ? | ? | ? | ? | ? |
| Household infrastructure, assets, ownership | Water source; toilet type/shared; cooking fuel; household assets; bank account; floor/roof/walls; house/land/item ownership; sleeping rooms |  | ? |  |  | ? |  |  | ? | ? | ? | ? |  |
| Household food security | Preferred foods unavailable; fewer meals; skipped meals |  | ? |  | ? |  | ? |  | ? |  | ? |  | ? |
| Index pregnancy | Date of last menstrual period; weeks pregnant; pregnancy wantedness/intentions and partner agreement; mental health symptoms during pregnancy; perceived impacts of pregnancy; legal indications for abortion (rape/incest and provider-identified maternal/fetal risk) | ? |  |  |  |  |  |  |  |  |  |  |  |
| History of contraceptive use | Ever used contraception; contraceptive use at time of conception; contraceptive decision-making (respondent/husband/joint) |  | ? |  |  |  |  |  |  |  |  |  |  |
| Health seeking at enrollment facility | Travel time/mode/cost; lodging; barriers; accompaniment; decision-making; primary reason for abortion | ? |  |  |  |  |  |  |  |  |  |  |  |
| Abortion care seeking before enrollment | Prior attempts to end pregnancy (number; method; people and providers involved; pill type/route; settings; care; reasons for no care) | ? |  |  |  |  |  |  |  |  |  |  |  |
| Abortion at enrollment facility: care quality & outcomes | Method of abortion; counseling (use, expected symptoms, complications); regimen/dose/route; pill ID; completion of abortion; complications; post-abortion care and treatment; post-abortion physical limitations |  | ? |  |  |  |  |  |  |  |  |  |  |
| Denied abortion at enrollment facility: Initial post-denial pathways | Reason and context for denial; details of subsequent abortion attempts (methods, sources, costs, travel, lodging, complications); current pregnancy status; details of miscarriage (if relevant) |  | ? |  |  |  |  |  |  |  |  |  |  |
| Denied abortion at enrollment facility: Later post-denial pathways<sup>c</sup> | Pregnancy outcome (miscarriage/abortion/birth/still pregnant); details of subsequent abortion attempts (methods, sources, costs, travel, lodging, complications, post-abortion functional limitations); details of miscarriage (if relevant) |  |  | ? |  |  |  |  |  |  |  |  |  |
| Subsequent pregnancies | Any new pregnancies since last interview; attempts to end subsequent pregnancies (method, people involved, pills/regimen/route, settings, care/treatment, costs, complications, post-abortion physical limitations); current pregnancy status; date of last menstrual period and weeks pregnant (if currently pregnant); subsequent pregnancy outcomes (miscarriage, live birth, stillbirth) |  |  | ? | ? | ? | ? | ? | ? | ? | ? | ? | ? |
| Knowledge of abortion legality | General legality; specific circumstances; guardian/husband permission |  | ? |  |  |  |  |  |  |  |  |  |  |
| Maternal perinatal health<sup>d</sup> | health provider and visits during pregnancy; maternal health and nutrition during pregnancy; sex determination; delivery place/attendant; delivery payments; postpartum maternal complications; post-birth physical limitations |  | ? | ? | ? | ? | ? | ? | ? | ? | ? | ? | ? |
| Infant health<sup>d</sup> | Live or stillbirth; infant death (cause); child sex; birth weight/size; preterm; breastfeeding practices |  | ? | ? | ? | ? | ? | ? | ? | ? | ? | ? | ? |
| Mother–infant bonding<sup>e</sup> |  |  | ? | ? | ? | ? | ? | ? | ? | ? | ? | ? | ? |
| Child preventive care & recent illness<sup>f</sup> | Vaccinations; vitamin A/iron/deworming; diarrhea (symptoms and care); fever/cough (symptoms and care); disability; serious injuries |  | ? | ? | ? | ? | ? | ? | ? | ? | ? | ? | ? |
| Age-specific child development screening<sup>f</sup> |  |  | ? | ? | ? | ? | ? | ? | ? | ? | ? | ? | ? |
| Women's empowerment and relationship quality | Household decision-making power; mobility/freedom, relationship happiness, husband alcohol consumption | ? |  |  | ? |  | ? |  | ? |  | ? |  | ? |
| Relationship commitment/trust |  |  | ? |  | ? |  | ? |  | ? |  | ? |  | ? |
| Violence<sup>g</sup> | Psychological, economic, physical and sexual violence from intimate partner; violence from non-partner(s) | ? |  |  | ? |  | ? |  | ? |  | ? |  | ? |
| Mental health screening<sup>h</sup> | Generalized anxiety disorder; depression; post-traumatic stress disorder; stress; life satisfaction; community abortion stigma | ? | ? | ? | ? | ? | ? | ? | ? | ? | ? | ? | ? |
| Physical health—self-rating & diagnoses<sup>i</sup> | Self-rated health; clinician-diagnosed conditions (anemia, hypertension, diabetes, cholesterol, asthma; chronic respiratory disease, cancer); types of chronic pain | ? | ? | ? | ? | ? | ? | ? | ? | ? | ? | ? | ? |
| Fertility intentions & contraceptive use<sup>j</sup> | Current pregnancy intentions/wantedness; recent contraceptive use; reasons for nonuse |  | ? | ? | ? | ? | ? | ? | ? | ? | ? | ? | ? |
| Anthropometry<sup>k</sup> | Weight, height, and mid upper arm circumference |  | ? | ? | ? | ? | ? | ? | ? | ? | ? | ? | ? |

Notes: *BL*=baseline; *w*=weeks, *m*=months.
a. Limited respondent demographics recorded at 6w if already taken at BL, time-invariant demographics asked only once (BL or 6w)
b. Husband demographics only asked after 6w if newly married since prior wave
c. Later post-denial pathways asked at 6m for those still pregnant at 6w, asked at first follow-up visit for participants that were unreachable at 6w
d. Maternal perinatal health and infant health indicators measured for index and subsequent pregnancies ending in births only
e. Maternal-infant bonding measured for all children child <18 months (index, subsequent, or prior children)
f. Child preventive care & recent illness and child development measured for all children < 3 years (index, subsequent, and prior children)
g. Sexual violence not asked at 12m or 24m
h. Abortion stigma asked only at 6w and at all follow-ups; Lifetime history of mental health conditions also recorded at BL
i. Both pre-pregnancy and current self-rated health measured at BL; Chronic respiratory disease and cancer only measured at follow-ups
j. Current fertility intentions & contraceptive use only asked if respondent reports not pregnant and not sterilized
k. Anthropometry measured for respondent and all children <3 years (index, subsequent, or prior children)

#### Definition of study groups

This cohort is unique in capturing abortion care seeking experiences and outcomes. All participants had one of four possible experiences with their index pregnancy: (1) received abortion at initial care seeking; (2) denied abortion initially but obtained one later; (3) denied abortion but later reported being no longer pregnant; or (4) denied abortion and subsequently gave birth. Initial abortion receipt or denial was determined via “yes” or “no” answers to the six-week survey question: “Did you have an abortion at [recruitment facility] on or around [baseline date]?” Those who did not complete the six-week survey because they were unavailable for interview were asked the same question at their first follow-up survey. Initially denied individuals were then asked about subsequent abortion attempts and their current pregnancy status, at all follow-up surveys until they reported being no longer pregnant or giving birth.

For future analyses, we propose that abortion denial be defined using two possible indicators:

(1) a three-arm indicator—initially received abortion (“Abortion” group), initially denied but did not carry the pregnancy to term (“Turnaway – No Birth” group), and denied abortion and gave birth (“Turnaway – Birth group); and (2) a binary indicator—”No Birth” group vs. “Gave Birth” group. Given the dearth of evidence on abortion seeking in this setting, there are reasonable justifications for both including those who were initially denied but did not carry the pregnancy to term as a separate exposure group and for grouping them in with those who initially received their abortions.

### Balancing weights

This study was modeled after the US Turnaway Study, in which abortion seekers nearing the gestational limit (and therefore left without other termination options if denied) were similar in most characteristics and only differed by whether they received an abortion, allowing for a natural experiment design to assess the effect of carrying an unwanted pregnancy to term versus receiving an abortion over time. In the Nepal setting however, we observed that abortion denial was dependent on factors in addition to gestational age, including provider availability, beliefs and bias, and individual sociodemographic characteristics.[19] Further differentiation in who was able to end their pregnancy occurred because many of those who were initially denied an abortion did not carry the pregnancy to term and give birth and, among those denied, pregnancy outcomes differed by sociodemographic characteristics.

Given the systematic differences by abortion denial and pregnancy outcome, we created balancing weights to match covariate distributions across the exposure groups. Exposure-based weighting is preferred over standard regression adjustment (another option to strengthen causal inferences) given the study’s focus on multiple health and economic effects of abortion denial and the large number of potential confounders for these analyses, which would each require correct specification in all models. To ensure consistency across planned outcome analyses and facilitate use of this data by potential collaborators, we constructed two sets of balancing weights to allow for analyses using either the binary exposure (“Gave Birth” vs. “No Birth”) or the three-arm exposure (“Abortion” vs. “Turnaway – No Birth” vs. “Turnaway – Birth”).

Propensity scores were estimated using multivariable regression models predicting pregnancy outcome. To support analyses using a binary pregnancy outcome exposure, multivariable logistic regression was used, while multinomial logistic regression was applied for weights that could be used in three-arm exposure analyses. Predictors in propensity score models included the following variables measured at baseline: sociodemographic characteristics (age, marital status, education, employment, parity, income), pregnancy characteristics (gestational age, abortion seeking due to fetal anomaly or sex selection, pregnancy intendedness), clinic characteristics (distance from participant home, public/private), medical history (anxiety/depression, chronic pain, overall health, intimate partner violence), and measures of empowerment and husband drinking behaviors. These predictors of pregnancy outcome were determined based on subject-matter expertise from the study team. Propensity scores were then used to generate overlap weights, which are calculated using an approach similar to inverse probability of treatment weighting. These weights, when applied to the full sample, create a pseudo-population in which baseline characteristics are balanced between exposure groups, facilitating estimation of causal effects.[20] We compared baseline demographic characteristics across pregnancy outcomes before and after applying the weights. Covariate balance was evaluated using standardized mean differences. Additional details on the construction of weights and diagnostic assessments are provided in S1 File.

Data processing and description was conducted using conducted using Stata SE version 17.0 (StataCorp, College Station, TX). Statistical analyses and assessment of weight performance were conducted using R version 4.4.2 (R Core Team, Vienna, Austria).

## RESULTS

### Participation

Between April 2019 and December 2020, 8,849 women sought abortion care at the 22 recruitment sites in this study (Fig 2). 1,916 (22%) were eligible for the study and 1,832 (96% of eligible women) agreed to participate and complete a baseline interview at the recruitment facility. Nine individuals initially identified as pregnant at recruitment were later determined to have never been pregnant and excluded from analyses. At the six-week follow-up, these individuals reported not having received an abortion due to one of the following reasons: a negative pregnancy test or ultrasound, return of menstruation, or self-reporting that they were not pregnant. The criteria for excluding these individuals was finalized after publication of baseline data from this cohort,[19] and thus final baseline sample size presented here are slightly lower than those in the study’s first publication (1,832 vs 1,835).

**Figure 2.**
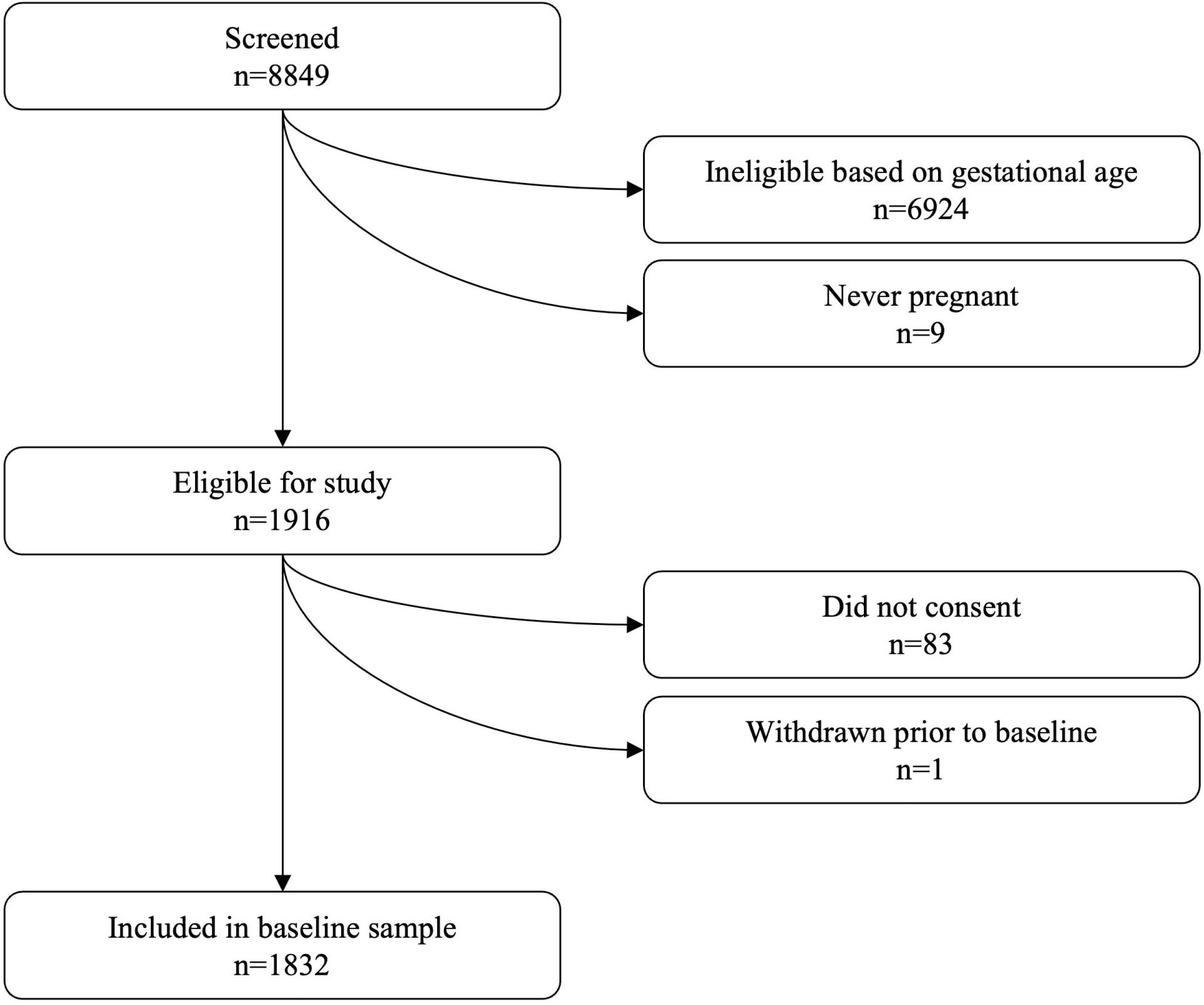
Study recruitment.

### Pregnancy outcomes

Of the 1,832 women who completed baseline surveys, 86 were lost before the six-week survey and four were lost after completing the six-week follow-up survey, at which point they were still pregnant (Fig 3). These 90 women (less than 5% of the baseline sample) were excluded from analyses due to the unknown outcome of their pregnancy, resulting in an analytical sample of 1742 women. Compared to participants with known pregnancy outcomes, these 90 women were more likely to be younger, unmarried, of Dalit ethnicity, and at greater than 10 weeks’ gestation or uncertain gestation at baseline. They were also more likely to have fewer children and to report lower pregnancy wantedness for the index pregnancy.

**Figure 3.**
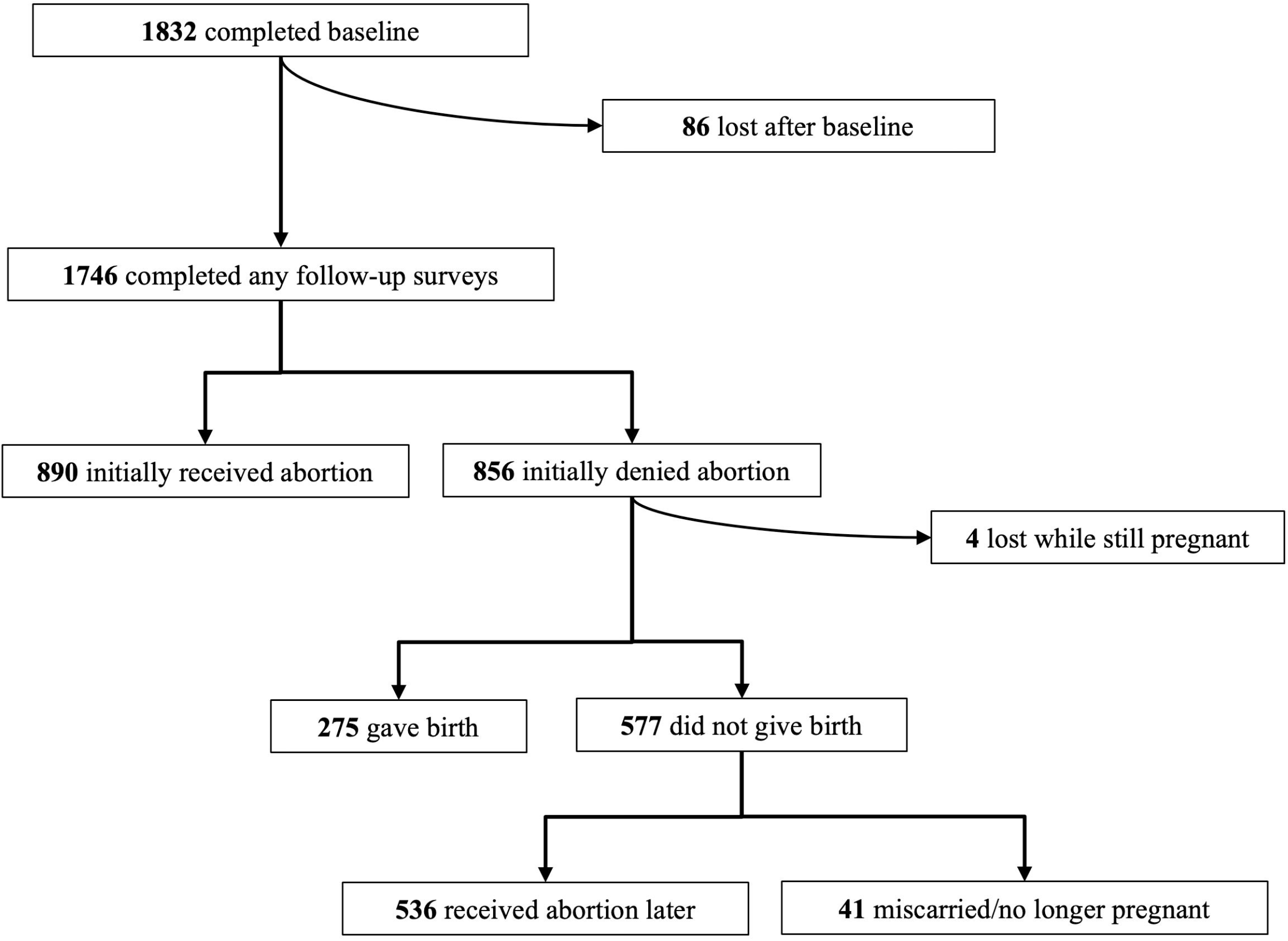
Classification of pregnancy outcomes.

Of the 1,742 individuals with follow-up data, 890 received an abortion at that facility shortly after the time of recruitment, while 856 were initially denied. Of those denied, 275 ultimately gave birth. However, subsequent abortion attempts were reported by the majority of those initially denied, with 536 reporting later receiving an abortion. An additional 41 participants reported that they were initially denied abortion but were no longer pregnant. Of these, 29 reported no subsequent abortion attempts and that they had a miscarriage, nine did not specify how the pregnancy ended, and three reported having a miscarriage following a subsequent abortion attempt. There is ambiguity around these cases, but we classified them according to the participant’s chosen language around their pregnancy outcome and all were ultimately categorized into the No Birth category. Four individuals who reported being still pregnant at the six-week survey after initial denial were lost to follow-up and excluded from any analyses that examined outcomes based on pregnancy outcome. Some pregnancy outcomes were ascertained after the publication of preliminary baseline data from this cohort, leading to slight differences in the analytical sample size (1,742 vs 1,663) presented here, compared to the study’s first publication.[19]

### Retention

Participants were followed for an average of 56 months (standard deviation = 12.4). Participation by survey round is show in Table 2. Assessments of retention are complicated by our ceasing of data collection in December 2024 when funding was exhausted. Only participants recruited prior to December 2019 were able to complete the full 60 months of interviews; all others were administratively censored. By the time of study completion, 1073 participants (62%) had completed 60-month interviews; 478 (27%) were still actively participating but were administratively censored after completing the 48-month or 54-month survey; 191 (11%) had been previously lost to follow-up.

**Table 2.** Number of interviews and retention by survey round.

| **Table 2. Number of interviews and retention by survey round** |  |  |
| --- | --- | --- |
|  | **No. interviews** | **% retained since baseline** |
| **baseline** | 1,832 | -- |
| **6 week** | 1,665 | 91 |
| **6 month** | 1,644 | 90 |
| **12 month** | 1,631 | 89 |
| **18 month** | 1,645 | 90 |
| **24 month** | 1,648 | 90 |
| **30 month** | 1,656 | 90 |
| **36 month** | 1,642 | 90 |
| **42 month** | 1,580 | 86 |
| **48 month** | 1,550 | 85 |
| **54 month** | 1,303 | 71 |
| **60 month** | 1,057 | 58 |

\*Start of administrative censoring

Participants who received their abortion have on average three months longer follow-up than those who carried to term (56.5 vs. 53.7). This is a consequence of the fact that in the first month of recruitment, everyone seeking abortion was eligible for the study regardless of gestation. After the first month, recruitment was limited to those later in pregnancy (ten or more weeks) and those who did not know their gestational duration. Those recruited in the first month were both less likely to be denied an abortion or give birth (because they had earlier gestations on average) and more likely to complete 60 months of follow-up. Those recruited in subsequent months skewed later in pregnancy and were more likely to be denied an abortion and to give birth, as well as to be administratively censored by the end of data collection. Given that abortion denial differed by baseline characteristics, differences were also present across administrative censoring groups for the following indicators: caste, education, employment, facility type, clinic travel distance, gestational age, empowerment, income, and reason for abortion seeking (sex selection or fetal anomaly). However, as this informative censoring was administrative and dependent wholly on gestational age, outcomes for these individuals can be classified as “Missing at Random”, meaning missingness is entirely dependent on known and measured variables.[21] Correctly specified analysis models using maximum-likelihood-based methods remain robust to informative loss-to-follow-up in cases where the “Missing at Random” assumptions are satisfied.[21] Thus, as long as analysis models for this study account for gestational age (either as a covariate in regression adjustment or as part of the balancing weights), and employ maximum-likelihood-based methods (including linear and generalized mixed models), it is reasonable to assume that all estimates are unbiased by administrative censoring.

Eleven percent of participants (n=191) were lost to follow-up after their pregnancy outcome was ascertained. After accounting for clustering by facility, loss to follow-up was nondifferential with respect to pregnancy outcome. Among the 1,095 participants not at risk for administrative censoring (ie. recruited early enough to complete all 60 months of follow-up, 63% of all respondents), there was no association between pregnancy outcome and attrition, either in two-group analyses (No Birth: 11.6% vs Gave Birth: 11.8%; p=0.965), or three-group analyses (Abortion: 12.0% vs Turnaway – No Birth: 10.5% vs Turnaway – Birth: 11.8% p=0.784).

Additionally, no systematic differences in characteristics were observed among those lost early compared to those retained to the end of the study.

### Cohort characteristics at baseline

A total of 1742 women were retained in the study long enough to ascertain their pregnancy outcome and are used as the analysis dataset for this cohort. Table 3 contains overall, unweighted sample characteristics. Almost all women (97%, n=1694) were married at baseline, 33% (579) were under age 25, 29% (n=504) were 25-29 years, 22% (n=380) were 30-34 years, and 16% (n=279) were over age 35. Nearly two-fifths belonged to the Brahmin/Chhetri/Thakuri castes (39%, n=683), followed by Hill Janajati (24%, n=415), Terai Janajati (22%, n=378), and Dalit or other (15%, n=259). The majority (68%, n=1188) completed secondary or higher education, while 16% (n=279) had no schooling and 15% (n=267) had completed primary school. Just over half of participants were working at baseline (54%, n=935) and 74% (n=1270) reported adequate or more than adequate household income. Most women had at least one child (87%, n=1511) and 58% had at least one son (n=1016). Approximately 10% sought abortions for reasons of sex selection or fetal anomaly (n=167 and n=181, respectively). 37% (n=648) were at or below ten weeks a time of abortion seeking, 37% were above ten weeks (n=635), and 26% (n=459) did not know their gestational age. Nearly three quarters sought abortion services at private facilities (71%, n=1244), and 46% (n=787) traveled over an hour to reach the facility.

**Table 3.** Distribution of baseline characteristics by two-arm exposure, unweighted and weighted.

|  | Unweighted |  |  |  | Weighted |  |  |
| --- | --- | --- | --- | --- | --- | --- | --- |
|  | Total<br>N=1742 | No Birth<br>N=1467 | Gave<br>Birth<br>N=275 | Standardi<br>zed mean<br>difference | No Birth | Gave<br>Birth | Standardized<br>mean<br>difference |
| **Demographic characteristics** |  |  |  |  |  |  |  |
| Age, years |  |  |  |  |  |  |  |
| ≤24 | 579 (33.2) | 469 (32.0) | 110 (40.0) | 0.17 | (38.6) | (38.6) | <0.01 |
| 25-29 | 504 (28.9) | 415 (28.3) | 89 (32.4) | 0.09 | (32.6) | (32.6) | <0.01 |
| 30-34 | 380 (21.8) | 332 (22.6) | 48 (17.5) | 0.13 | (17.8) | (17.8) | <0.01 |
| 35+ | 279 (16.0) | 251 (17.1) | 28 (10.2) | 0.20 | (10.9) | (10.9) | <0.01 |
| Marital status |  |  |  |  |  |  |  |
| Unmarried | 48 (2.8) | 42 (2.9) | 6 (2.2) | -- | (2.6) | (2.6) | -- |
| Married | 1694 (97.2) | 1425 (97.1) | 269 (97.8) | 0.04 | (97.4) | (97.4) | <0.01 |
| Caste |  |  |  |  |  |  |  |
| Brahmin/Chhetri/Thakuri | 683 (39.4) | 594 (40.6) | 89 (32.6) | 0.16 | (34.5) | (34.2) | <0.01 |
| Hill Janajati | 415 (23.9) | 346 (23.7) | 69 (25.3) | 0.04 | (24.9) | (25.1) | <0.01 |
| Dalit/other | 259 (14.9) | 192 (13.1) | 67 (24.5) | 0.29 | (21.6) | (21.7) | <0.01 |
| Terai Janajati | 378 (21.8) | 330 (22.6) | 48 (17.6) | 0.13 | (19.0) | (19.1) | <0.01 |
| Education |  |  |  |  |  |  |  |
| None/ some formal schooling | 279 (16.1) | 230 (15.7) | 49 (17.9) | 0.06 | (17.4) | (17.6) | <0.01 |
| Primary | 267 (15.4) | 203 (13.9) | 64 (23.4) | 0.24 | (21.1) | (21.1) | <0.01 |
| Secondary | 1069 (61.6) | 920 (63.0) | 149 (54.6) | 0.17 | (56.9) | (56.6) | <0.01 |
| Higher | 119 (6.9) | 108 (7.4) | 11 (4.0) | 0.15 | (4.7) | (4.7) | <0.01 |
| Employment status |  |  |  |  |  |  |  |
| Not currently working | 800 (46.1) | 658 (45.0) | 142 (52.0) | -- | (51.1) | (50.8) | -- |
| Currently working | 935 (53.9) | 804 (55.0) | 131 (48.0) | 0.14 | (48.9) | (49.2) | <0.01 |
| No. of living children |  |  |  |  |  |  |  |
| 0 | 231 (13.3) | 200 (13.6) | 31 (11.3) | 0.07 | (12.1) | (12.1) | <0.01 |
| 1 | 520 (29.9) | 438 (29.9) | 82 (29.8) | <0.01 | (30.0) | (30.0) | <0.01 |
| 2 | 621 (35.6) | 530 (36.1) | 91 (33.1) | 0.06 | (33.8) | (33.8) | <0.01 |
| 3+ | 370 (21.2) | 299 (20.4) | 71 (25.8) | 0.13 | (24.1) | (24.1) | <0.01 |
| Sex of living children |  |  |  |  |  |  |  |
| No boys | 726 (41.7) | 608 (41.4) | 118 (42.9) | -- | (43.9) | (43.9) | -- |
| 1+ boys | 1016 (58.3) | 859 (58.6) | 157 (57.1) | 0.03 | (56.1) | (56.1) | <0.01 |
| Income adequacy |  |  |  |  |  |  |  |
| Not Adequate | 463 (26.7) | 357 (24.4) | 106 (39.0) | 0.31 | (34.7) | (35.0) | <0.01 |
| Adequate | 1228 (70.9) | 1066 (73.0) | 162 (59.6) | 0.28 | (63.7) | (63.4) | <0.01 |
| More than adequate | 42 (2.4) | 38 (2.6) | 4 (1.5) | 0.08 | (1.5) | (1.5) | <0.01 |
| **Pregnancy and clinic factors** |  |  |  |  |  |  |  |
| LMUP Score | 3.7 (2.7) | 3.7 (2.7) | 3.8 (2.6) | 0.05 | 3.7 (2.6) | 3.7 (2.6) | <0.01 |
| Seeking for sex selection | 167 (9.6) | 116 (7.9) | 51 (18.5) | 0.32 | (16.5) | (16.5) | <0.01 |
| Seeking for fetal anomaly | 181 (10.4) | 174 (11.9) | 7 (2.5) | 0.37 | (3.4) | (3.4) | <0.01 |
| Gestational age |  |  |  |  |  |  |  |
| at or below 10 weeks | 648 (37.2) | 613 (41.8) | 35 (12.7) | 0.69 | (16.3) | (16.3) | <0.01 |
| above 10 weeks | 635 (36.5) | 486 (33.1) | 149 (54.2) | 0.43 | (51.0) | (51.0) | <0.01 |
| don't know gestation | 459 (26.3) | 368 (25.1) | 91 (33.1) | 0.18 | (32.6) | (32.6) | <0.01 |
| Seeking at private facility | 1244 (71.4) | 1047 (71.4) | 197 (71.6) | 0.01 | (72.3) | (72.3) | <0.01 |
| Travel distance to facility |  |  |  |  |  |  |  |
| Less than 30 minutes | 535 (31.2) | 482 (33.3) | 53 (19.6) | 0.31 | (21.8) | (21.6) | <0.01 |
| 30 minutes to 1 hour | 395 (23.0) | 331 (22.9) | 64 (23.7) | 0.01 | (24.4) | (24.8) | <0.01 |
| 1 to 3 hours | 413 (24.1) | 333 (23.0) | 80 (29.6) | 0.16 | (28.2) | (27.8) | <0.01 |
| 3+ hours | 374 (21.8) | 301 (20.8) | 73 (27.0) | 0.14 | (25.6) | (25.8) | <0.01 |
| **Health and empowerment factors** |  |  |  |  |  |  |  |
| Chronic pain |  |  |  |  |  |  |  |
| No | 996 (57.4) | 830 (56.8) | 166 (60.6) | 0.08 | (59.7) | (59.6) | <0.01 |
| Yes | 684 (39.4) | 582 (39.8) | 102 (37.2) | 0.05 | (37.5) | (37.6) | <0.01 |
| Don't know | 56 (3.2) | 50 (3.4) | 6 (2.2) | 0.07 | (2.8) | (2.8) | <0.01 |
| Pre-pregnancy self-rated health |  |  |  |  |  |  |  |
| Good or very good | 1575 (90.7) | 1333 (91.2) | 242 (88.3) | -- | (88.9) | (88.9) | -- |
| Poor or very poor | 161 (9.3) | 129 (8.8) | 32 (11.7) | 0.09 | (11.1) | (11.1) | <0.01 |
| History of anxiety or depression | 551 (31.7) | 459 (31.4) | 92 (33.6) | 0.05 | (31.9) | (32.0) | <0.01 |
| Husband's alcohol consumption |  |  |  |  |  |  |  |
| Never or rarely | 982 (58.2) | 836 (59.0) | 146 (54.5) | 0.10 | (56.2) | (56.3) | <0.01 |
| 1-2x per month | 356 (21.1) | 308 (21.7) | 48 (17.9) | 0.09 | (18.5) | (18.5) | <0.01 |
| At least 1x per week | 192 (11.4) | 157 (11.1) | 35 (13.1) | 0.06 | (12.0) | (12.0) | <0.01 |
| Every day | 156 (9.3) | 117 (8.3) | 39 (14.6) | 0.20 | (13.3) | (13.2) | <0.01 |
| Intimate partner violence | 388 (23.0) | 313 (22.1) | 75 (28.0) | 0.14 | (25.1) | (25.0) | <0.01 |
| Limited mobility | 413 (23.7) | 329 (22.4) | 84 (30.5) | 0.18 | (28.2) | (28.2) | <0.01 |
| Household decision-making power |  |  |  |  |  |  |  |
| None | 866 (51.4) | 707 (49.9) | 159 (59.3) | 0.18 | (58.4) | (58.5) | <0.01 |
| Some | 244 (14.5) | 212 (15.0) | 32 (11.9) | 0.08 | (12.0) | (12.0) | <0.01 |
| More | 575 (34.1) | 498 (35.1) | 77 (28.7) | 0.13 | (29.6) | (29.5) | <0.01 |

Notes: Unweighted data are presented as N(column %) for categorical measures and mean(SD) for ordinal measures. Exposure balancing weights derived from propensity scores predicting exposure (gave birth). Weighted results apply to artificially balanced sample and thus counts are not shown.

Most women rated their pre-pregnancy health as good or very good (91%, n=1575), though 39% (n=684) reported a history of chronic pain, 32% (n=551) reported a lifetime history of anxiety or depression and 23% (n=388) experienced intimate partner violence. About half (51%, n= 866) lacked household decision-making power, 24% (n=413) reported that their husband limited their mobility outside the home, and 9% (n=156) reported that their husband drank alcohol daily.

### Baseline differences by study group and weight performance

The data in Table 3 demonstrate that those who gave birth differed at baseline from those who did not give birth. Those who gave birth were more likely to be younger, less likely to belong to the Brahmin, Chhetri, or Thakuri castes, and more likely to be in the disadvantaged Dalit caste; they were less likely to have secondary or higher education, less likely to be employed outside the home, and more likely to report having an inadequate income. They were also less likely to live within half an hour of the abortion facility. Those who gave birth were much more likely to have been at or above ten weeks of pregnancy than those who receive an abortion and they were more likely to be seeking abortion for reasons of sex selection but less likely to be seeking abortion for fetal anomaly. Women who gave birth were more likely to have a husband who drinks daily, have experienced intimate partner violence, lack decision-making power in the household, and to have limited mobility outside the home. Standardized mean differences greater than 0.1 are reflective of statistically significant differences.[22]

Table 4 similarly shows differences in baseline characteristics among the three study groups. In several cases, the two turnaway groups resembled each other but differed from those who received an abortion upon initial care seeking: both were more likely than the abortion group to present above ten weeks’ gestation and to have no living children. In other cases, the Turnaway–Birth group stood apart from the other two groups who did not give birth, being more likely to be younger than 25 years but less likely to be over 35 years, less likely to be Brahmin/Chhetri/Thakuri, more likely to be Dalit/other caste, less likely to have secondary or higher education, and more likely to have a husband who drinks daily.

**Table 4.** Distribution of baseline sociodemographic characteristics by three-arm exposure, unweighted and weighted.

|  |  |  |  |  |  |  |  |  |  |
| --- | --- | --- | --- | --- | --- | --- | --- | --- | --- |
|  | Unweighted |  |  |  |  | Weighted |  |  |  |
|  | Total<br>N=1742 | Abortion<br>n=890 | Turnaway-No Birth<br>n=577 | Turnaway -<br>Birth<br>n=275 | Standardized<br>mean<br>difference | Abortion | Turnaway<br>-<br>No Birth | Turnaway-Birth | Standardized<br>mean<br>difference |
| **Demographic characteristics** |  |  |  |  |  |  |  |  |  |
| Age, years |  |  |  |  |  |  |  |  |  |
| ≤24 | 579 (33.2) | 290 (32.6) | 179 (31.0) | 110 (40.0) | 0.19 | (35.1) | (37.8) | (36.8) | 0.06 |
| 25-29 | 504 (28.9) | 244 (27.4) | 171 (29.6) | 89 (32.4) | 0.11 | (33.0) | (31.5) | (32.5) | 0.03 |
| 30-34 | 380 (21.8) | 207 (23.3) | 125 (21.7) | 48 (17.5) | 0.14 | (19.5) | (18.2) | (18.4) | 0.03 |
| 35+ | 279 (16.0) | 149 (16.7) | 102 (17.7) | 28 (10.2) | 0.21 | (12.4) | (12.6) | (12.3) | 0.01 |
| Marital status |  |  |  |  |  |  |  |  |  |
| Unmarried | 48 (2.8) | 17 (1.9) | 25 (4.3) | 6 (2.2) | -- | (2.9) | (3.2) | (2.9) | -- |
| Married | 1694 (97.2) | 873 (98.1) | 552 (95.7) | 269 (97.8) | 0.15 | (97.1) | (96.8) | (97.1) | 0.02 |
| Caste |  |  |  |  |  |  |  |  |  |
| Brahmin/Chhetri/Thakuri | 683 (39.4) | 352 (39.7) | 242 (42.0) | 89 (32.6) | 0.19 | (38.1) | (36.5) | (35.4) | 0.05 |
| Hill Janajati | 415 (23.9) | 236 (26.6) | 110 (19.1) | 69 (25.3) | 0.18 | (24.7) | (25.8) | (24.9) | 0.03 |
| Dalit/other | 259 (14.9) | 116 (13.1) | 76 (13.2) | 67 (24.5) | 0.31 | (18.7) | (19.0) | (19.7) | 0.02 |
| Terai Janajati | 378 (21.8) | 182 (20.5) | 148 (25.7) | 48 (17.6) | 0.20 | (18.4) | (18.6) | (20.0) | 0.04 |
| Education |  |  |  |  |  |  |  |  |  |
| None/ some formal schooling | 279 (16.1) | 132 (14.9) | 98 (17.0) | 49 (17.9) | 0.08 | (17.8) | (16.4) | (17.3) | 0.05 |
| Primary | 267 (15.4) | 130 (14.7) | 73 (12.7) | 64 (23.4) | 0.29 | (18.4) | (18.0) | (19.9) | 0.05 |
| Secondary | 1069 (61.6) | 563 (63.6) | 357 (62.0) | 149 (54.6) | 0.18 | (58.2) | (61.3) | (57.8) | 0.07 |
| Higher | 119 (6.9) | 60 (6.8) | 48 (8.3) | 11 (4.0) | 0.18 | (5.6) | (4.2) | (5.0) | 0.05 |
| Employment status |  |  |  |  |  |  |  |  |  |
| Not currently working | 800 (46.1) | 390 (44.0) | 268 (46.5) | 142 (52.0) | -- | (49.5) | (48.3) | (49.0) | -- |
| Currently working | 935 (53.9) | 496 (56.0) | 308 (53.5) | 131 (48.0) | 0.16 | (50.5) | (51.7) | (51.0) | 0.03 |
| No. of living children |  |  |  |  |  |  |  |  |  |
| 0 | 231 (13.3) | 137 (15.4) | 63 (10.9) | 31 (11.3) | 0.14 | (11.7) | (12.6) | (12.9) | 0.04 |
| 1 | 520 (29.9) | 279 (31.3) | 159 (27.6) | 82 (29.8) | 0.08 | (31.8) | (31.0) | (31.0) | 0.02 |
| 2 | 621 (35.6) | 304 (34.2) | 226 (39.2) | 91 (33.1) | 0.13 | (32.9) | (33.3) | (33.1) | 0.01 |
| 3+ | 370 (21.2) | 170 (19.1) | 129 (22.4) | 71 (25.8) | 0.16 | (23.6) | (23.1) | (22.9) | 0.01 |
| Sex of living children |  |  |  |  |  |  |  |  |  |
| No boys | 726 (41.7) | 352 (39.6) | 256 (44.4) | 118 (42.9) | -- | (39.0) | (40.4) | (42.2) | -- |
| 1+ boys | 1016 (58.3) | 538 (60.4) | 321 (55.6) | 157 (57.1) | 0.10 | (61.0) | (59.6) | (57.8) | 0.06 |
| Income adequacy |  |  |  |  |  |  |  |  |  |
| Not Adequate | 463 (26.7) | 191 (21.6) | 166 (28.9) | 106 (39.0) | 0.38 | (31.2) | (32.5) | (32.0) | 0.03 |
| Adequate | 1228 (70.9) | 670 (75.6) | 396 (68.9) | 162 (59.6) | 0.34 | (67.7) | (65.4) | (66.4) | 0.05 |
| More than adequate | 42 (2.4) | 25 (2.8) | 13 (2.3) | 4 (1.5) | 0.09 | (1.1) | (2.1) | (1.6) | 0.07 |
| **Pregnancy and clinic factors** |  |  |  |  |  |  |  |  |  |
| LMUP Score | 3.7 (2.7) | 3.6 (2.6) | 3.8 (2.8) | 3.8 (2.6) | 0.08 | 3.5 (2.6) | 3.5 (2.6) | 3.5 (2.4) | 0.02 |
| Seeking for sex selection | 167 (9.6) | 27 (3.0) | 89 (15.4) | 51 (18.5) | 0.48 | (10.9) | (10.1) | (11.0) | 0.03 |
| Seeking for fetal anomaly | 181 (10.4) | 130 (14.6) | 44 (7.6) | 7 (2.5) | 0.45 | (4.5) | (4.9) | (5.5) | 0.04 |
| Gestational age |  |  |  |  |  |  |  |  |  |
| at or below 10 weeks | 648 (37.2) | 453 (50.9) | 160 (27.7) | 35 (12.7) | 0.88 | (22.2) | (20.3) | (21.7) | 0.04 |
| above 10 weeks | 635 (36.5) | 193 (21.7) | 293 (50.8) | 149 (54.2) | 0.69 | (44.3) | (45.1) | (45.4) | 0.02 |
| don't know gestation | 459 (26.3) | 244 (27.4) | 124 (21.5) | 91 (33.1) | 0.26 | (33.5) | (34.6) | (32.9) | 0.04 |
| Seeking at private facility | 1244 (71.4) | 584 (65.6) | 463 (80.2) | 197 (71.6) | 0.33 | (73.5) | (71.9) | (73.1) | 0.04 |
| Travel distance to facility |  |  |  |  |  |  |  |  |  |
| Less than 30 minutes | 535 (31.2) | 318 (36.2) | 164 (28.9) | 53 (19.6) | 0.36 | (26.9) | (25.2) | (24.4) | 0.04 |
| 30 minutes to 1 hour | 395 (23.0) | 197 (22.4) | 134 (23.6) | 64 (23.7) | 0.03 | (22.8) | (23.2) | (25.7) | 0.05 |
| 1 to 3 hours | 413 (24.1) | 199 (22.6) | 134 (23.6) | 80 (29.6) | 0.17 | (25.6) | (26.1) | (24.8) | 0.02 |
| 3+ hours | 374 (21.8) | 165 (18.8) | 136 (23.9) | 73 (27.0) | 0.19 | (24.7) | (25.5) | (25.2) | 0.02 |
| **Health and empowerment factors** |  |  |  |  |  |  |  |  |  |
| Chronic pain |  |  |  |  |  |  |  |  |  |
| No | 996 (57.4) | 489 (55.2) | 341 (59.2) | 166 (60.6) | 0.11 | (58.2) | (59.9) | (58.9) | 0.03 |
| Yes | 684 (39.4) | 378 (42.7) | 204 (35.4) | 102 (37.2) | 0.15 | (38.8) | (37.8) | (37.6) | 0.02 |
| Don't know | 56 (3.2) | 19 (2.1) | 31 (5.4) | 6 (2.2) | 0.18 | (3.0) | (2.3) | (3.5) | 0.07 |
| Pre-pregnancy self-rated health |  |  |  |  |  |  |  |  |  |
| Good or very good | 1575 (90.7) | 805 (90.9) | 528 (91.7) | 242 (88.3) | -- | (88.8) | (90.4) | (88.5) | -- |
| Poor or very poor | 161 (9.3) | 81 (9.1) | 48 (8.3) | 32 (11.7) | 0.11 | (11.2) | (9.6) | (11.5) | 0.06 |
| History of anxiety or depression | 551 (31.7) | 269 (30.4) | 190 (33.0) | 92 (33.6) | 0.07 | (31.9) | (33.0) | (31.0) | 0.05 |
| Husband's alcohol consumption |  |  |  |  |  |  |  |  |  |
| Never or rarely | 982 (58.2) | 503 (58.0) | 333 (60.4) | 146 (54.5) | 0.13 | (59.5) | (58.9) | (57.1) | 0.05 |
| 1-2x per month | 356 (21.1) | 187 (21.6) | 121 (22.0) | 48 (17.9) | 0.09 | (18.6) | (17.7) | (19.5) | 0.04 |
| At least 1x per week | 192 (11.4) | 100 (11.5) | 57 (10.3) | 35 (13.1) | 0.09 | (10.7) | (12.4) | (11.0) | 0.05 |
| Every day | 156 (9.3) | 77 (8.9) | 40 (7.3) | 39 (14.6) | 0.24 | (11.2) | (10.9) | (12.4) | 0.05 |
| Intimate partner violence | 388 (23.0) | 209 (24.1) | 104 (18.9) | 75 (28.0) | 0.22 | (24.2) | (24.0) | (24.4) | 0.01 |
| Limited mobility | 413 (23.7) | 187 (21.0) | 142 (24.6) | 84 (30.5) | 0.22 | (28.1) | (25.5) | (26.0) | 0.06 |
| Household decision-making power |  |  |  |  |  |  |  |  |  |
| None | 866 (51.4) | 410 (47.3) | 297 (53.9) | 159 (59.3) | 0.23 | (56.7) | (53.8) | (57.1) | 0.06 |
| Some | 244 (14.5) | 139 (16.1) | 73 (13.2) | 32 (11.9) | 0.12 | (11.7) | (13.7) | (12.3) | 0.06 |
| More | 575 (34.1) | 317 (36.6) | 181 (32.8) | 77 (28.7) | 0.16 | (31.6) | (32.6) | (30.6) | 0.04 |

*Notes:* Unweighted data are presented as N(column %) for categorical measures and mean(SD) for ordinal measures. Exposure balancing weights derived from propensity scores predicting exposure (abortion denial/pregnancy outcome). Weighted results apply to artificially balanced sample and thus counts are not shown. Standardized mean difference for multiarm abortion denial exposure calculated using minimum and maximum means from the three study groups.

As shown in Table 3 (and Fig C in S1 File), applying exposure balancing weights from propensity score models eliminated all baseline differences across the two abortion exposure groups. This occurs because the “overlap” weighting method employed in this study mathematically guarantees perfect covariate balance specifically when there are only two groups.[23] In contrast, as shown in Table 4 (and Fig D in S1 File), weights did not produce perfect balance with three exposure categories. Detailed formulas for weight generation are provided in S1 File. Nonetheless, for the three-arm abortion exposure, the weights achieved adequate covariate overlap, with all mean absolute standardized differences below 0.1.[22] Further assessment of weight performance and validity was confirmed by additional balance diagnostics, found in Fig A and Fig B in S1 File.

### Additional findings to date

Baseline data from the Nepal Turnaway Study were previously used to describe the characteristics of abortion seekers overall and by care received, the burden and context of legal abortion denial, and care seeking trajectories following denial.[19] Puri and colleagues found that most who were denied (84%) were told that they were too far along, although many should have been deemed legally eligible for abortion upon this first care seeking attempt according to current laws. The most common reasons for seeking abortion focused on existing family structures – already having enough children, having a very young child, and being unable to afford another child.

Albach and colleagues investigated prior abortion attempts in this cohort [24], finding that 14% of participants attempted to end their pregnancy before enrolling, with 83% using abortion medication and 79% obtaining it from pharmacists. Women who struggled to access abortion clinics were more likely to rely on pharmacy-dispensed medication, emphasizing the critical role of pharmacists in abortion care provision.

A COVID-19 analysis from the Nepal Turnaway Study cohort [25] revealed that 93% of participants experienced pandemic-related impacts, with economic challenges, difficulty accessing necessities, and interruptions to children’s education being common. Women denied abortion and later giving birth faced greater difficulties accessing health services and financial challenges, suggesting the pandemic worsened social and health inequities, including barriers to abortion access.

### STRENGTHS AND LIMITATIONS

This cohort allows researchers to study the effects of abortion denial in a context with high maternal mortality, documented practice of abortion outside the certified medical system, and high poverty levels. We followed abortion seekers to observe whether they were denied their abortion, their eventual pregnancy outcome, and their subsequent social, health, and economic wellbeing. Most of the studies that have attempted to estimate the morbidity that results from unintended pregnancies or unwanted births rely on cross-sectional or retrospective data.[26] Retrospective studies suffer from potentially severe biases. Given the stigma associated with abortion, women are often unwilling to report past abortions – or they report them as miscarriages – leading to significant selection bias.[27,28] The Nepal Turnaway Study recruited individuals prior to the end of their pregnancy, enabling researchers to examine the predictors and consequences of the outcomes of unwanted pregnancy.

This paper proposes both two-arm and three-arm exposure definitions to assess the impact of different pregnancy outcomes. In both definitions, those who obtained a legal, facility-based abortion offer the appropriate counterfactual for both illegal abortion and carrying an unwanted pregnancy to term. Causal claims regarding the effects of abortion denial using data from this cohort are strengthened by the initiation of data collection prior to participants’ having been denied or received an abortion. This establishes temporality by guaranteeing that the outcomes observed did not precede the exposure. Additionally, recruitment of participants at the point of abortion seeking allows us to control for risk factors common to women with unwanted pregnancy who seek abortion (the target population). We used propensity score weighting based on more than 15 baseline characteristics and their interactions to improve covariate balance and allow researchers to estimate average treatment effects. Specifically, we applied overlap weighting, a technique which ensures that treatment effects are estimated among individuals with sufficient representation across study groups. This approach is particularly useful when certain subgroups are highly likely to fall into only one exposure arm-- for example, participants presenting at the earliest possible gestation were very likely to obtain an abortion. By prioritizing individuals who could reasonably fall within all exposure groups, overlap weights yield more precise estimates of average treatment effects relevant to real-world situations.

In addition to collecting data on the health and wellbeing of women after being denied or obtaining an abortion, this study also followed the children of these women, both those born of an unwanted pregnancy and women’s existing children at the time of abortion seeking. The majority of women seeking abortion services in this setting are already mothers and cite desire to care for one’s existing children is a common reason for seeking an abortion.[19] Yet little data on the future outcomes of these existing children are available. Finally, in Nepal where societal attitudes about gender cultivate son preference,[29] data on the outcomes of girl children born following denial of abortion services are important to understand and inform policy around sex-selective abortion.

This study has several limitations to consider for future analyses. First, propensity score weighting accounts only for measured confounders. Unmeasured or unknown factors that influence both whether someone is denied an abortion and subsequent outcomes may still introduce residual confounding. To the extent that such factors are strongly correlated with included covariates, they may be partially or fully accounted for. Second, several potential sources exist for selection bias. While less than five percent of individuals were lost to follow-up after baseline, these women lack pregnancy outcome data and differ from those retained in several key characteristics. Additionally, 27% of respondents included in longitudinal analyses were administratively censored after 48-54 months of follow-up (instead of followed through 60 months), limiting the power of analyses that include the last year of follow-up. However, this administrative censoring was caused by intentional changes in recruitment strategy aimed at selecting individuals with later gestation and thus is completely explained by baseline characteristics incorporated into models. In this situation, estimates derived from maximum-likelihood-based methods are relatively unbiased by the presence of censoring [16]. Lastly, 11% of respondents were lost to follow-up after their pregnancy outcome was ascertained, although this attrition was nondifferential with respect to abortion denial.

## CONCLUSIONS

The Nepal Turnaway Study is the first prospective cohort to examine the experiences and trajectories of abortion seekers and their families in this context. Longitudinal data collected at the point of abortion receipt or denial make it possible to examine outcomes among women who were denied but did not carry to term, as well as those who experienced unwanted childbirth, compared with women who obtained legal procedures. Exposure balancing weights developed for this cohort will allow future researchers to isolate the health and economic impacts of abortion denial while accounting for factors associated with unwanted pregnancy. In addition, this study considers a wide range of consequences, including not only women’s physical health and economic outcomes, but also their mental and emotional well-being and the health of their children.

## Data Availability

Upon publication, data will be publicly available for five years at a third party website. To gain access, data requestors will need to sign a data access agreement.

## Acknowledgements

We thank the members of the National Technical Advisory Committee members including Dr. Bibek Kumar Lal, for their guidance and support. We are grateful to seven government hospitals, MSI, the Family Planning Association of Nepal, and the Sunaulo Bhabishya Nepal clinics for permitting us to conduct this study in their facilities and assisting with eligibility form completion. We acknowledge Preeti Gautam for project management support and Sara Daniels for data infrastructure. We thank the 24 research assistants for their dedicated work in data collection and, most importantly, the study participants for generously sharing their time and information.

## Collaboration and data sharing

The authors welcome proposals for collaboration, which should be directed to. The cohort dataset is available for five years from XXX database (accession number: XXX).

