## Supplementary material for "Cohort profile: The Nepal Turnaway Study": S1 File (Supplemental Material)

**S1 File. Construction and evaluation of balancing weights**

For two-arm exposure analyses, propensity scores were estimated using multivariable logistic regression. For three-arm exposure analyses, they were estimated using multivariable multinomial logistic regression. Both models included the following variables, selected based on the study team’s conceptual and empirical understanding of the predictors of abortion denial: age (≤24 years, 25-29 years, 30-34 years, 35+ years; marital status (binary), caste (Brahmin/Chhetri/Thakuri, Hill Janajati, Terai Janajati, Dalit/other), educational attainment (none/some formal schooling, primary, secondary, higher), employment status (binary), number of existing children (none, one, two, three or more), sex of existing children (any boys or no boys), self-reported household income (not adequate, adequate, more than adequate), distance traveled to facility(sample quartiles), facility type (private, public), gestational age (at or below ten weeks, above ten weeks, don’t know gestation), abortion seeking due to fetal anomaly (binary), abortion seeking due to sex selection (binary), self-rated physical health prior to pregnancy (poor/very poor vs. good/very good), lifetime history of anxiety or depression (binary), recent history of intimate partner violence (binary), husband’s drinking behaviors (never/rarely, 1-2x per month, at least one time per week, every day), (decision-making score (zero, one, two), mobility score (zero, one) and recent chronic pain (yes, no, don’t know).

We checked for the presence of nonlinear relationships between continuous variables (age, travel distance to clinic, and LMUP) and abortion denial through graphical and statistical tests for linear, quadratic, and cubic trends. Model fit was also evaluated by sequentially adding these terms to the propensity score model, retaining those that significantly improved fit (p<.05 for test H_0_: β= 0). Restricted cubic spline terms for these variables were explored but did not improve fit for either model. A categorical variable was chosen for gestation given that the recruitment strategy prioritized those greater than ten weeks and with an unknown gestation (27% of all participants). We also tested hypothesis-driven interactions, including caste by education, caste by age, gestational age by anxiety/depression history, and intimate partner violence by mobility score. While both caste by age and caste by education terms improved model fit in the two-arm and three-arm propensity models, the latter was excluded from the two-arm model due to positivity violations (empty cells for certain categories of caste crossed with education).

Mode imputation was performed for predictor variables with fewer than ten missing observations (<0.5% of the sample), including caste, education, employment, depression/anxiety history, chronic pain, pre-pregnancy health, and household income. Travel distance to the clinic (n=25 missing) was imputed to the site-specific median. Missing responses on intimate partner violence (n=57), decision-making (n=58), and husband drinking behaviors (n=57) were imputed to the mode. Sensitivity analyses excluding these cases from the propensity scores and subsequent analyses yielded consistent results.

Propensity scores were converted to weights to estimate the average treatment effect (ATE) in the “overlap” group, a subsample of individuals that have similar propensity scores across exposure groups.[20] Overlap weighting, a form of balancing weighting, yields results similar to those generated using inverse probability of treatment weighting (IPW), which estimates the ATE in the full population. In observational studies, ATE estimation can be challenging due to limited covariate overlap between exposure groups. Overlap weighting improves specificity without requiring outlier weight trimming, which is often necessary to deal with limited covariate overlap. Additionally, overlap weights are constrained between zero and one, leading to more precise estimates than traditional IPW. With overlap weighting for a two-arm exposure, each observation is assigned a weight based on the probability of being in the other exposure group:

$$\left\{ \begin{aligned} 1 - e(x), for Z = 1 \\ e(x), for Z = 0 \end{aligned} \right.$$

Where e(x) represents the predicted probability of abortion denial from the propensity score model, and Z=1 indicates that an individual gave birth and Z=0 indicates they received their abortion.

Overlap weighting can be generalized for analyses using a multi-arm exposure using the following formula:

$$\frac{\frac{1}{e_{j}(x)}}{\sum_{l=1}^{K} \frac{1}{e_{l}(x)}}$$

Where *j* represents each of the three abortion denial arms, e*_j_*(x) represents the predicted probability of an individual’s observed denial category, *j*, and $\sum_{l=1}^{K} \frac{1}{e_{l}(x)}$ represents the sum of an individual’s inverse predicted probabilities of falling into each of the three denial categories (*l*=1…*K*).[2] Under this generalization, each observation is assigned a weight based on *both* the probability of being in their observed exposure group, and probability of receiving all other exposures:

$$\left\{ \begin{aligned} \frac{\frac{1}{e_{1}\left( x \right)}}{\frac{1}{e_{1}\left( x \right)}+\frac{1}{e_{2}\left( x \right)}+\frac{1}{e_{3}\left( x \right)}} , for Z = 1 \\ \frac{\frac{1}{e_{2}\left( x \right)}}{\frac{1}{e_{1}\left( x \right)}+\frac{1}{e_{2}\left( x \right)}+\frac{1}{e_{3}\left( x \right)}} , for Z = 2 \\ \frac{\frac{1}{e_{3}\left( x \right)}}{\frac{1}{e_{1}\left( x \right)}+\frac{1}{e_{2}\left( x \right)}+\frac{1}{e_{3}\left( x \right)}} , for Z = 3 \end{aligned} \right.$$

Covariate balance was assessed via absolute standardized mean differences (shown in Fig A and Fig B) defined as:

$\Delta=\frac{\left| \bar{x}_{Z=1}-\bar{x}_{Z=0} \right|}{\sqrt{\frac{s_{Z=1}^{2}+ s_{Z=0}^{2}}{2}}} \mathrm{fo}$ for continuous covariates and $\Delta=\frac{\left| \hat{p}_{Z=1}-\hat{p}_{Z=0} \right|}{\sqrt{\frac{\hat{p}_{Z=1}(1-\hat{p}_{Z=1}) + \hat{p}_{Z=0}(1-\hat{p}_{Z=0})}{2}}}$ for binary covariates,

where $\left| \bar{x}_{Z=1}-\bar{x}_{Z=0} \right|$ is the absolute difference in the sample mean for a continuous covariate (exposed minus unexposed), $s_{Z=1}^{2}- s_{Z=0}^{2}$ is the difference in the sample variance for a continuous covariate, and $\left| \hat{p}_{Z=1}-\hat{p}_{Z=0} \right|$ is the absolute difference in the sample prevalence for a binary covariate. For the three-arm weights, differences were calculated using the highest and lowest mean values (or proportions) for each covariate. A maximum mean absolute standardized difference of 0.1 was used as a cutoff to ensure adequate covariate balance.[3]

Validity of the overlap weighting methods was also assessed by visually examining the distribution of propensity scores across study groups, to ensure sufficient covariate overlap. Covariate overlap was sufficient for the two-arm exposure (Fig A) and for predicting both Abortion and Turnaway-No Birth in the three-arm exposure (Fig B). Slightly less covariate overlap was present for predicting Turnaway-Birth in the three-arm exposure, particularly between the Turnaway-Birth and the Abortion groups. However, this is to be expected with this type of exposure, given that many of those who received their abortion upon initial attempt would be highly unlikely to experience the counterfactual of denial and giving birth based on their characteristics (gestational age, fetal indication etc.). A benefit of employing overlap weights is that the propensity scores do not need to be arbitrarily trimmed to eliminate extreme values, as the target marginal estimate is the treatment effect in the overlap group, not the full population.

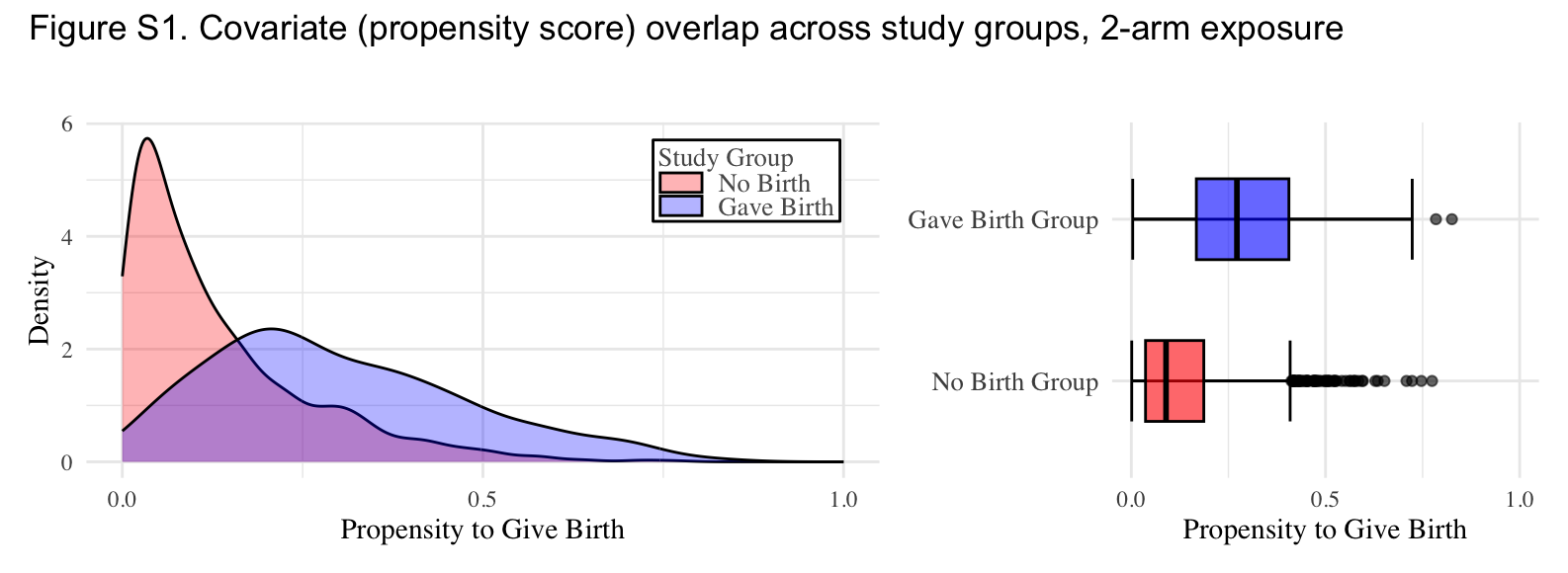

**Fig A. Covariate (propensity score) overlap across study groups, 2-arm exposure**

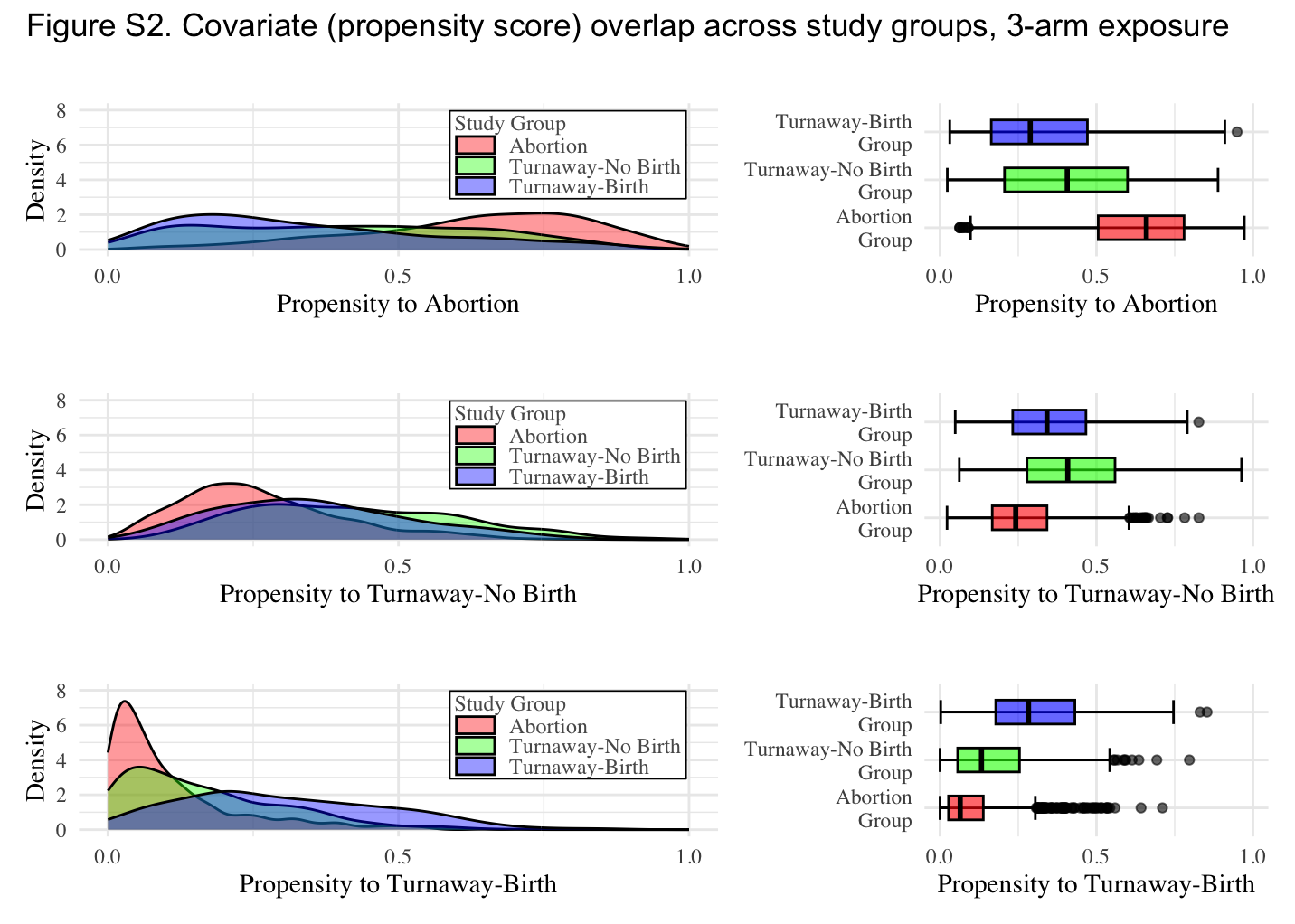

**Fig B. Covariate (propensity score) overlap across study groups, 3-arm exposure**

**Fig C. Covariate balance across study groups, 2-arm exposure**

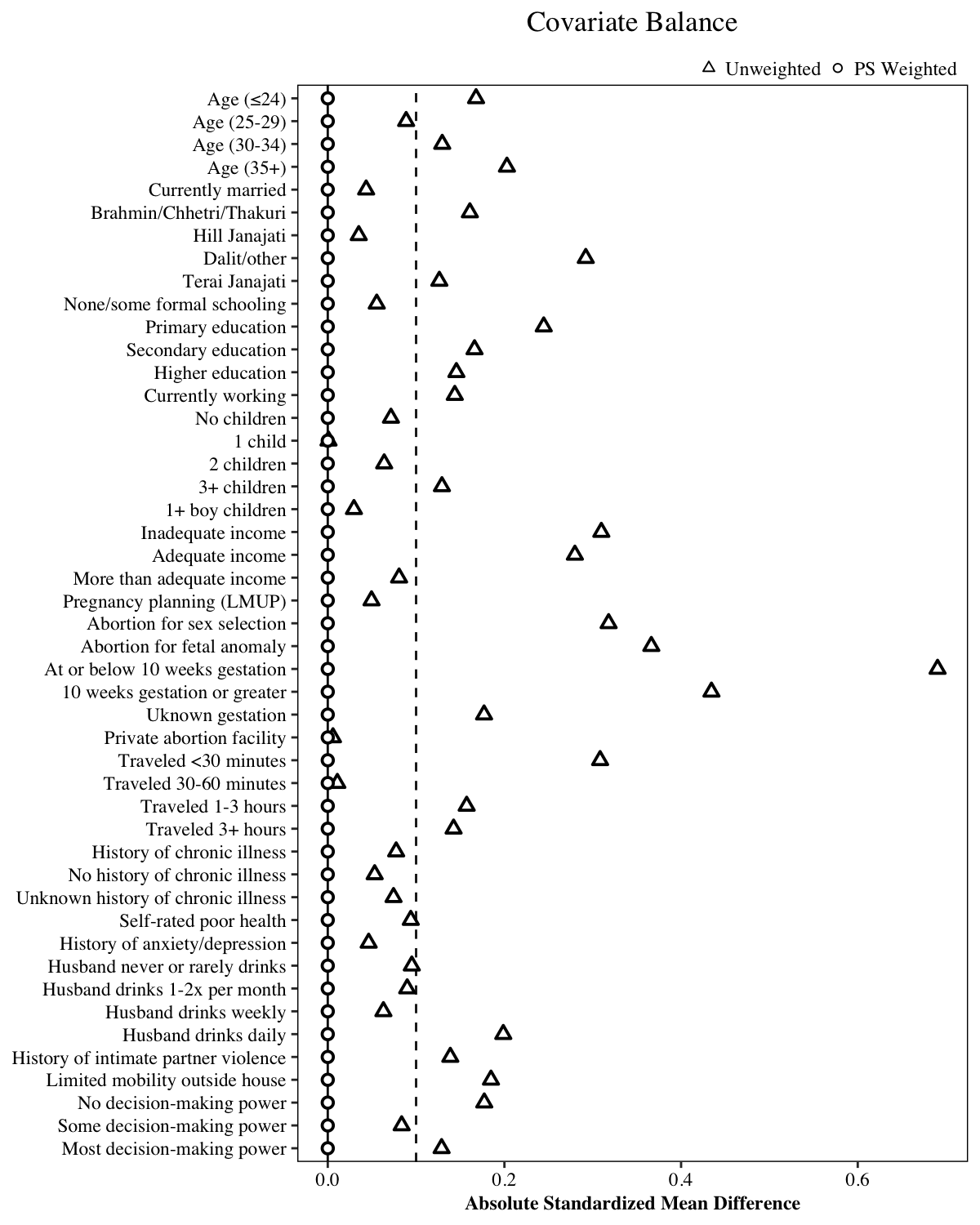

**Fig D. Covariate balance across study groups, 3-arm exposure**

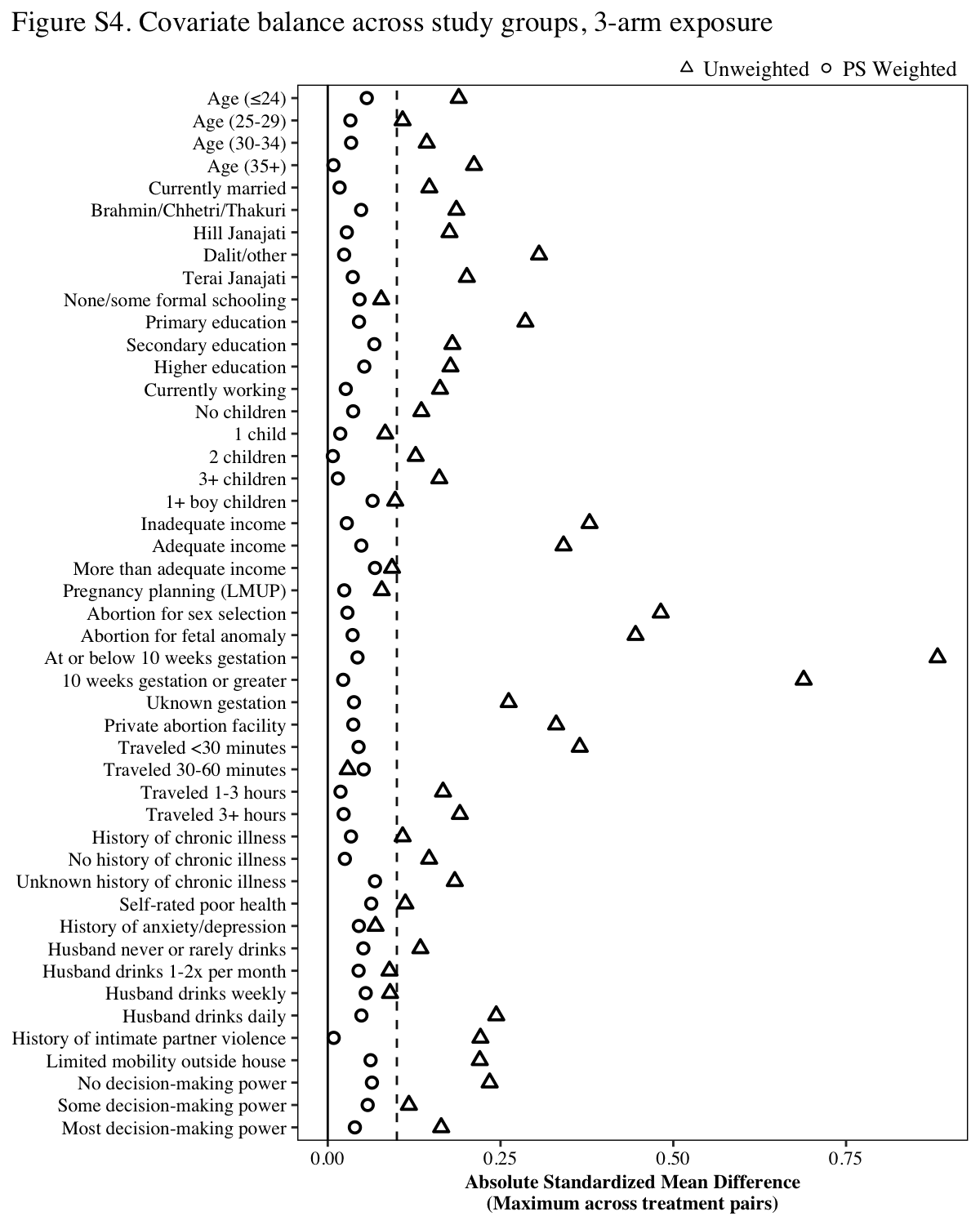
